# Validation of smartphone-based assessments of depressive symptoms using the Remote Monitoring Application in Psychiatry (ReMAP)

**DOI:** 10.1101/2020.08.27.20183418

**Authors:** Janik Goltermann, Daniel Emden, Elisabeth J. Leehr, Katharina Dohm, Ronny Redlich, Udo Dannlowski, Tim Hahn, Nils Opel

## Abstract

Smartphone-based symptom monitoring has gained increased attention in psychiatric research as a cost-efficient tool for prospective and ecologically valid assessments based on participants’ self-reports. However, a meaningful interpretation of smartphone-based assessments requires knowledge on their psychometric properties; especially their validity. Here, we conducted a systematic investigation of the validity of smartphone-based assessments of affective symptoms by using the smartphone app *Remote Monitoring Application in Psychiatry* (ReMAP). Beck Depression Inventory (BDI), and single-item mood and sleep information was assessed via the ReMAP app and validated with stationary (non-smartphone) BDI scores and clinician-rated depression severity using the Hamilton Depression Rating Scale (HDRS).

We found overall high comparability between smartphone-based and stationary BDI scores (ICC=.921, p<.001, n=173). Smartphone-based BDI further correlated with stationary HDRS ratings of depression severity (r=.783, p<.001, n=51). Higher agreement between smartphone and stationary assessments was found in affective disorder patients as compared to healthy controls, and anxiety disorder patients. Highly comparable agreement between delivery formats was found across age and gender groups. Similarly, smartphone-based single-item self-ratings of mood correlated with BDI sum scores (r=-.538, p<.001, n=168), while smartphone-based single-item sleep duration correlated with the sleep item of the BDI (r=-.310, p<.001, n=166).

The present findings demonstrate that smartphone-based monitoring of depressive symptoms via the ReMAP app provides valid assessments of depressive symptomatology and therefore represents a useful tool for prospective digital phenotyping in affective disorder patients in clinical and research applications.

## Introduction

The phasic development of symptoms over time in form of disease episodes is one of the key characteristics of affective disorders. These disease trajectories can be used as an informative predictor as well as an outcome measure in psychiatric research and personalized medicine. However, the assessment of the development of symptoms over time is challenging. The value of cross-sectional assessments is limited as they can only capture an excerpt of the symptom history and it is unclear whether this excerpt reflects e.g. the peak of an affective episode or a (partially) remitted state, and whether episodes are recurrent. Collecting this information retrospectively from the patients is one approach of gaining insights into the former symptom history, which albeit is likely to be biased by the current depressive state (Coyne, Thompson, & Racioppo, 2001). Thus, multiple prospective assessments of symptoms are needed for a valid interpolation to the underlying disease trajectory. Although such prospective instruments based on a paper-pencil format exist (Denicoff et al., 1997), their use is limited due to low cost-efficiency. In the recent years, the utilization of smartphone applications for psychological and psychiatric assessment have increased considerably due to their cost-efficiency and practicability (Onnela & Rauch, 2016; Russell & Gajos, 2020; Torous et al., 2018).

Several proof of concept studies have pointed to the utility of smartphone-based data in affective disorder research (Dogan et al., 2017). The potential of continuous monitoring of psychomotor activity based on acceleration and location for a differentiation of unipolar and bipolar patients has been demonstrated (Faurholt-Jepsen et al., 2012, 2015). Recent studies have also indicated that smartphone-based movement parameters allow for a prediction of intraindividual daily mood state changes (Faherty et al., 2017; Grünerbl et al., 2015; Pratap et al., 2019; Rohani, Faurholt-jepsen, Kessing, & Bardram, 2018; Torous et al., 2018). However, such prospective investigations require indepth knowledge of the psychometric properties of the acquired data especially when it comes to the validity of smartphone-based measurements. This point appears particularly important in study designs that entirely rely on smartphone-based data.

Consequently, the comparability between smartphone-based and stationary (conventional paper-pencil or stationary computer-based) versions of psychometric instruments have also received increasing attention (Dubad, Winsper, Meyer, Livanou, & Marwaha, 2018). Besides the obvious difference in the format in which content is presented, differences in the assessment setting (laboratory or clinical setting vs. variable situations in real-life), as well as technical reservations could lead to different assessment results. Particularly using smartphones, potential distractions may become more likely, with the environments of reporting participants being less controllable. Initial evidence suggests that scores derived from digital and paper-pencil psychometric instruments seem to be generally comparable, however with considerable variance in the agreement (Alfonsson, Maathz, & Hursti, 2014; van Ballegooijen, Riper, Cuijpers, van Oppen, & Smit, 2016). Yet, a considerable number of previous studies investigating the reliability and validity of digital phenotyping methods has focused on stationary computer-based assessments that might differ from mobile assessments via the participants’ smartphones as outlined above. For the Beck Depression Inventory interformat reliability between stationary paper-pencil and computer based versions has been demonstrated across several studies (Alfonsson et al., 2014), while large-scale validation reports of agreement between stationary and smartphone-based versions is currently lacking.

Data from pilot studies indicate agreement between smartphone-delivered self-rated mood and clinician-rated mood via Hamilton Depression Rating Scale (HDRS) scores in bipolar patients (Faurholt-Jepsen et al., 2014), and Juengst et al. demonstrated high comparability between mood related symptoms in traumatic brain injury patients assessed either via smartphone or via telephone interview (Juengst et al., 2015). In a systematic review of the literature including data from three studies and a total of n=89 bipolar outpatients, significant medium sized correlations between smartphone-based assessments of depressive symptoms and established clinical rating scales were reported (Faurholt-Jepsen, Munkholm, Frost, Bardram, & Kessing, 2016). Regarding smartphone-based monitoring in major depression, Torous et al. reported high agreement between smartphone-based and paper-pencil assessment using the PHQ-9 in 13 adult MDD patients (Torous et al., 2015), similarly Cao et al. reported agreement between smartphone-based self-reported mood and the PHQ-9 in n= 13 adolescent participants (Cao et al., 2020). One systematic review investigating the psychometric properties of mobile mood monitoring in young people concluded that there is enormous heterogeneity in the validity of smartphone-based delivery formats and more high-quality studies are needed (Dubad et al., 2018).

In sum, while the aforementioned findings of overall agreement between smartphone-based self-reported depressive symptoms and established clinical scales is encouraging, it appears important to denote that limited sample sizes of previous reports as well as systematic differences including sample properties, technical properties and assessment type currently limits our understanding of the reliability and validity of smartphone-based assessments of depressive symptoms. It thus remains unclear to which degree validation reports of smartphone-based self-reports are generalizable across assessment instruments, cohorts and applications and hence app- or study-specific validation of measurements remains the gold standard.

Therefore, the aim of the current study is to assess the validity of smartphone-based assessments of depressive symptoms using the ReMAP application. To this end, we investigate the comparability of smartphone-based self-reports of depressive symptoms with stationary (non-smartphone) versions of the Beck depression inventory (BDI), a well-established and standardized self-report instrument in psychiatric patients and healthy control participants. We test the hypotheses that both delivery formats – stationary and smartphone-based assessments – yield comparable results, and that hence smartphone-based monitoring of depressive symptoms via the ReMAP application provides valid assessments of depressive symptomatology. We furthermore aim to investigate potential differences in the agreement between smartphone- and stationary assessment of depressive symptoms across diagnostic groups, age groups and gender.

## Method

### Participants

The ReMAP study was designed as a prospective naturalistic observational study. An overall sample of N=173 participants is included in the current analyses. The sample includes adults that are either healthy controls (HC, n=101) or belong to one of the following diagnostic groups: MDD (n=43), bipolar disorder (BD, n=5), MDD with comorbid social anxiety disorder (SAD, n=9), SAD only (n=2), or specific phobia spider subtype (SP, n=13). Participants were recruited for ReMAP participation in the context of ongoing longitudinal cohort studies over which assessments were parallelized (details on subsamples from all cohorts are provided in the supplements).

Participants were informed about the possibility of a voluntary additional participation in the ReMAP study in a face-to-face meeting at the time they presented at the Department of Psychiatry in the context of ongoing longitudinal cohort assessments. Interested subjects were extensively briefed about aims, methods (especially type and amount of collected data), details on data security (details on data transfer and storage), and financial compensation. The study was approved by the local Institutional Review Board (IRB) and written informed consent was obtained before participation.

### Stationary Measures and procedures

All measures that are non-smartphone-based (conventionally administered in interviews or via paper-pencil or tablet questionnaires) will be referred to as “stationary assessments” and are described below. Presence or absence of a psychiatric diagnosis was assessed in all participants via a structured clinical interview for DSM-IV (SCID-I) (American Psychiatric Association, 2000; Wittchen, Wunderlich, Gruschwitz, & Zaudig, 1997) prior to participation in the ReMAP study. All healthy control participants were free from any history of a psychiatric disorder. As part of the original study assessments, participants from all cohorts provided self-reports of depressive symptoms via the BDI-I (Beck & Steer, 1987) or the BDI-II (Beck et al., 1996). Both Versions of the BDI are standardized and valid instruments for the assessments of depressive symptoms, and represent well-established assessment tools in research and clinical routine for the presence and extent of depressive symptoms. For the MACS and MNC cohorts additional assessments of clinician rated depression severity via the Hamilton Depression Rating Scale (HDRS) (Hamilton, 1960) were available.

### Smartphone App ReMAP

The Remote Monitoring Application in Psychiatry (ReMAP) is being developed at the Institute for Translational Psychiatry in Münster since mid-2018. It is a native app for iOS and Android, based on Apple ResearchKit, Apple Health, and Google Fit. After an anonymous login with a provided subject ID, the app works in background mode and monitors the number of steps taken by the user, the distance walked, the accelerometer, and GPS position data. The data is encrypted on the smartphone and sent regularly via REST-API to a backend specifically developed for ReMAP, which is provided on university servers. In addition, the app regularly enables the user to fill out various questionnaires regarding sleep and mood, as well as to create short voice recordings. Measures used in the current analyses are described below.

### Smartphone-based Measures and procedures

After written informed consent was obtained, each participant was provided an individual subject ID (subject code). The participant was then asked to download the developed smartphone app ReMAP and to start the application. At this time, subjects were asked to confirm again participation in the study and to enter the individual subject ID.

In addition to the continuous assessment of passive data, all participants were asked to provide self-reported ratings of depressive symptoms. To this end, participants were enabled to fill out a digital version of the BDI-I integrated in ReMAP every two weeks. Moreover, participants were enabled to rate their mood and sleep duration by answering single items every three days. For the single mood question (“How is your mood today?”) the participants provided their response via touch-screen on a scale from 1-10 with the anchors 1= “very bad” and 10= “very good”. For the single sleep question (“How many hours did you sleep last night?”) participants provided their response on a scale from 0 to 13 hours. For all self-reported data, the app sent out weekly push notifications on a random basis during daytime with variance of two days or every two weeks in case of BDI. Participants were instructed that answering to all questions is optional and they were free to choose their time of answering whenever items were made available.

For the present study, smartphone and stationary data were only included if the time interval between completions of the ratings between both delivery formats was smaller than 4 weeks to minimize potential bias due to temporal change in depressive symptoms. Further, for each participant the respective BDI, mood and sleep assessments from the time point with the shortest interval between smartphone-based and stationary assessment were included for the present study.

### Statistical analyses

Agreement between stationary and smartphone-administered BDI scores was assessed by absolute agreement in a two-way mixed-effects intraclass correlation (ICC) (Qin, Nelson, McLeod, Eremenco, & Coons, 2019). This analysis was further repeated for <1 week and the >1 week interval groups separately in order to assess the influence of the test-retest interval on the agreement between measurements. In addition, the analysis was repeated separately in healthy controls, affective disorder (MDD, SAD/MDD, and BD), and anxiety disorder (SP, SAD) patients as well as for the two stationary BDI versions (BDI-I and BDI-II). The internal consistency of the smartphone-based BDI was assessed via Cronbach’s alpha and compared with the internal consistency of the stationary BDIs.

For validation of the smartphone-based single mood item, it was correlated with the stationary and smartphone-based BDI scores. For validation of the smartphone-based single sleep item, it was correlated with the smartphone-based and stationary-administered BDI item assessing sleeping disturbance.

For further validation, the ReMAP BDI and the ReMAP single mood item were both correlated with clinician rated depression severity using the HDRS.

All analyses were conducted using SPSS (IBM, version 26), assuming a significance threshold of p<.05.

## Results

### Descriptive Statistics

Mean BDI scores and range across all participants were similar for ReMAP and stationary BDIs (mean=5.35, range 0-44; mean=6.46, range 0-47 respectively). Absolute differences between both measurements were on average 3.02 points with a considerable range covering 0-26 points. The mean test-retest interval was 5.84 days, ranging from 0.20 to 28.70 days. Detailed descriptive statistics over subgroups of the sample are provided in Table S1.

### Validity of affective symptom assessment via ReMAP

The overall agreement between ReMAP and stationary BDI was very high (ICC=.921, CI[.890, .942]). Separate investigations of the BDI agreement in several subgroups yielded highly comparable ICCs across both BDI versions (BDI-I and BDI-II), across different test-retest intervals, across different age groups, and across males and females (in all subgroups ICC>.888). Separate investigations across different diagnostic statuses yielded the highest BDI agreement between delivery formats in the subgroup with affective disorders (ICC=.912), while HCs and participants with anxiety disorders (SP and SAD) showed moderate agreement between BDIs (ICC=.639 and ICC=.736 respectively). ICC statistics for the full sample and all subgroups are presented in Table 1. Scatter plots of ReMAP BDI scores over stationary BDI scores are provided in Figure 1.

**Table 1.** Intraclass correlation agreement of ReMAP BDI-I with full sample and stratified subsamples.

|  | ICC | lower CI | upper CI | p | n |
| --- | --- | --- | --- | --- | --- |
| Full sample | .921 | .890 | .942 | <.001 | 173 |
| BDI-I<sub>stationary</sub> | .921 | .870 | .952 | <.001 | 64 |
| BDI-II<sub>stationary</sub> | .919 | .863 | .850 | <.001 | 109 |
| <1 week interval | .934 | .890 | .958 | <.001 | 126 |
| >1 week interval | .888 | .799 | .938 | <.001 | 47 |
| Healthy control | .639 | .454 | .760 | <.001 | 101 |
| Affective disorders | .912 | .851 | .948 | <.001 | 57 |
| Anxiety disorders | .736 | .252 | .910 | .008 | 15 |
| Age ≤35 | .899 | .851 | .931 | <.001 | 131 |
| Age >35 | .962 | .930 | .980 | <.001 | 42 |
| Male | .969 | .919 | .986 | <.001 | 41 |
| Female | .904 | .864 | .933 | <.001 | 132 |

**Figure 1.**
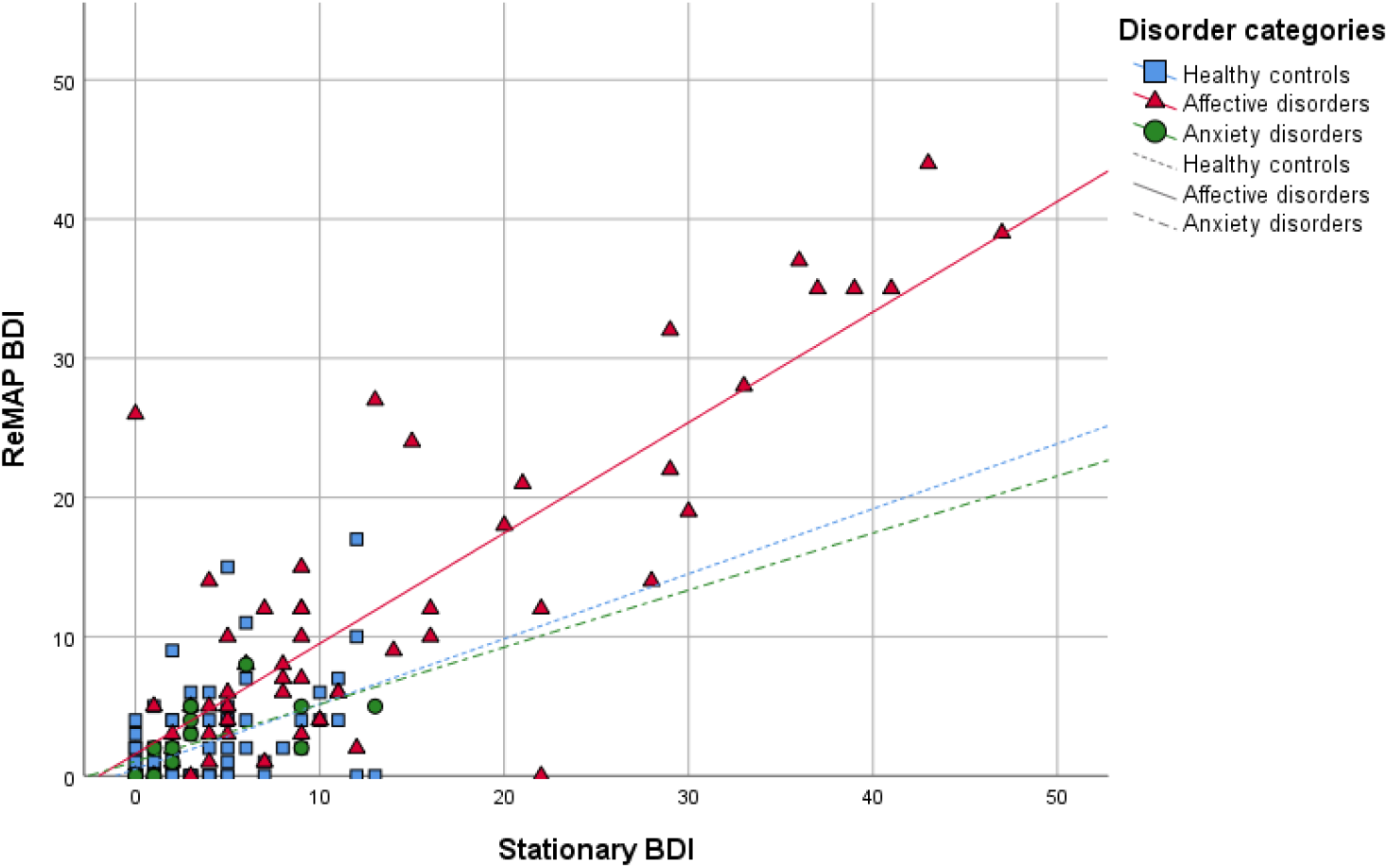
ReMAP smartphone BDI scores over stationary BDI scores across diagnostic groups. BDI, Beck Depression Inventory.

The internal consistency of the ReMAP BDI (Cronbach’s α=.944, n=174) was virtually identical to both stationary BDI versions (BDI-I: α=.945, n=54; BDI-II: α=.944, n=108).

For further validation the ReMAP BDI was correlated with clinician rated depression severity using the HDRS in a subset of the sample (n=51). The analysis yielded a strong significant correlation (r=.783, p<.001) that was comparable to the association between the HDRS score and the score of the stationary BDI (r=.682, p<.001).

Including all data points with a test-retest interval up to four weeks the single item for mood assessed via ReMAP correlated moderately with the sum scores of the ReMAP BDI (r=-.538, p<.001, n=168), and both stationary BDI versions (BDI-I: r=-.485, p<.001, n=61; BDI-II: r=-.504, p=<.001, n=107). Further, a significant negative correlation between ReMAP single mood item and HDRS score was observed (r=-.369, p=.008, n=51). Correlations of the single mood item over subsamples are provided in Table S2 in the supplements.

The single item for sleep assessed via ReMAP was correlated with the BDI item assessing sleeping disturbance. Including all data points with retest intervals up to four weeks, this analysis yielded significant negative associations with the sleep item from the ReMAP BDI (r=-.310, p<.001, n=166), and with the sleep item of both stationary BDI versions (BDI-I: r=-.279, p=.027, n=63; BDI-II: r=-.202, p=.042, n=102). Separate correlation analyses of the single mood and sleep ReMAP items across disorder subgroups are presented in Table S3 in the supplements. The general pattern of results yielded strongest associations in the affective disorder group.

## Discussion

With the present study we demonstrate that smartphone-based monitoring of depressive symptoms via the ReMAP app provides valid assessments of depressive symptomatology. The overall high agreement between the stationary and smartphone-based versions of the BDI confirm that digital assessments via the ReMAP application using the participants smartphone has the potential to offer valid estimates of the trajectory of participants mood. This notion is additionally supported by the observed correlation of smartphone-administered single-item ratings regarding mood and sleep with corresponding stationary assessments. Importantly, the validity of smartphone-based assessments could furthermore be demonstrated by using clinical rating scales as criterion with a strong correlation of smartphone-based BDI and stationary HDRS scores.

The observation of high agreement between self-reported smartphone-based assessments of depressive symptoms and classic stationary assessments in the present study is supported by previous findings from pilot studies in MDD patients (Torous et al., 2015) and from a systematic review in bipolar patients (Faurholt-Jepsen et al., 2016). Furthermore, the comparability of the stationary and smartphone version of the BDI in our study matches similar results of agreement between paper-pencil and stationary computer versions of the BDI (Alfonsson et al., 2014).

Our findings of overall high validity of smartphone-based and conventional stationary assessments of depressive symptoms in a relatively large and heterogeneous sample critically underscores the potential of mobile assessment tools in psychiatric research. Considering that smartphone-based assessments offer valid data on patients’ mood state, an expansion of mobile data acquisition in clinical and research context appears desirable. The cost-efficiency of smartphone-based data might thus allow to acquire valid data on patients’ long-term disease trajectory in an unprecedented scale. Together with previous studies investigating the comparability of BDI-I and BDI-II versions (Beck et al., 1996; Wang & Gorenstein, 2013), as well as delivery formats (Alfonsson et al., 2014), our findings add to an increasing evidence of high comparability of smartphone-based and conventional stationary assessments of depressive symptoms.

We furthermore demonstrate that agreement between smartphone-based and stationary assessment of depressive symptoms does not depend on participants’ age or gender supporting the generalizability of smartphone-based assessment of depressive symptoms. This notion appears especially noteworthy considering the relatively large sample size in comparison with previous reports as well as the age range of participants included in the current study (18 – 68 years).

An important observation of the present study was that higher agreement between smartphone and stationary assessments of depressive symptoms could be found in affective disorder patients compared to anxiety disorder patients or healthy controls – a finding that might at least partly be traced back to the much higher variability of depressive symptoms in the affective disorder group. Notably, while the agreement in the affective disorder sample can be estimated as excellent, intraclass correlations indicate a lower but still moderate to good agreement in the HC and anxiety disorder samples (Portney & Watkins, 2000). Nonetheless, these findings should be taken into account by future research and call for a more cautious interpretation of findings based on self-reported symptom data in healthy populations.

Besides validation of a smartphone version of the BDI, the present study furthermore found moderate to high agreement between mood ratings via smartphone-based single item assessments and established clinical scores using the BDI (regardless of the delivery format of the BDI). This finding is of particular importance considering that completion of an entire questionnaire is time-consuming and hence the usage of single items might provide a valid possibility of assessing mood on a frequent basis. Compared with the single mood item, the single sleep item showed a lower correlation with corresponding stationary assessments in the form of sleep disturbance items within the BDI questionnaire. One possible explanation for this finding could be that variability in the sleep quality is less temporally stable as compared to mood changes. Thus, the test interval of up to four weeks may be too long in order to validate the smartphone-based assessment of sleep quality. Considering that smartphone-based and stationary assessment methods lie several days or weeks apart, the association between them seems to be reasonably high. Further, sleep quality or disturbance may be a more heterogeneous construct and thus more difficult to assess via a single item.

Strengths of the present study comprise the relatively large sample of participants and the availability of smartphone-based along with conventional psychometric and clinical data. The present study furthermore included participants with differing psychiatric diagnoses and a high variability in age thus allowing to assess the generalizability across such participant groups. Further, a wide variety of assessments forms were used for validation, considering multiple sources of information. The application of stationary BDI versions (self-report), as well as clinical ratings (HDRS) underlines the validity of the smartphone-based assessments via the ReMAP app. Limitations include the lack of prospective clinical follow-up data. Future large-scale studies are warranted to assess the prognostic validity of smartphone-based self-reports in affective disorder patients.

Smartphone-based monitoring of depressive symptoms remains a timely matter of critical relevance for translational psychiatry. The present results demonstrate overall high validity of smartphone-based assessments of depressive symptoms and should thus encourage researchers to apply mobile applications for continuous prospective assessments of depressive symptoms.

## Data Availability

Derived data supporting the findings of this study are available from the corresponding author on request.

## Acknowledgements

EJL was supported by the Christiane Nüsslein-Vollhard Foundation.

Principal investigators in the FOR2107 consortium that are not co-authors of the current paper are: Work package (WP) 6, multi-method data analytics: Bertram Müller-Myhsok, Astrid Dempfle. Central project (CP) 1, biobank: Petra Pfefferle, Harald Renz. CP2, administration: Carsten Konrad.

Further acknowledgements:

Henrike Bröhl, Bruno Dietsche, Rozbeh Elahi, Jennifer Engelen, Sabine Fischer, Jessica Heinen, Svenja Klingel, Felicitas Meier, Torsten Sauder, Annette Tittmar, Dilara Yüksel (Dept. of Psychiatry, Marburg University). Mechthild Wallnig, Rita Werner (Core-Facility Brainimaging, Marburg University). Carmen Schade-Brittinger, Maik Hahmann (Coordinating Centre for Clinical Trials, Marburg). Michael Putzke (Psychiatric Hospital, Friedberg). Rolf Speier, Lutz Lenhard (Psychiatric Hospital, Haina). Birgit Köhnlein (Psychiatric Practice, Marburg). Peter Wulf, Jürgen Kleebach, Achim Becker (Psychiatric Hospital Hephata, Schwalmstadt-Treysa). Ruth Bär (Care facility Bischoff, Neukirchen). Matthias Müller, Michael Franz, Siegfried Scharmann, Anja Haag, Kristina Spenner, Ulrich Ohlenschläger (Psychiatric Hospital Vitos, Marburg). Matthias Müller, Michael Franz, Bernd Kundermann (Psychiatric Hospital Vitos, Gießen). Katharina Förster, Kordula Vorspohl, Bettina Walden, Dario Zaremba, Lena Waltemate, Dominik Grotegerd, Joscha Böhnlein, Tiana Borgers, Verena Enneking, Stella Fingas, Marius Gruber, Carina Hülsmann, Hannah Lemke, Susanne Meinert, Maike Richter, Lisa Sindermann, Katharina Thiel (Dept. of Psychiatry, University of Münster). Harald Kugel, Walter Heindel, Birgit Vahrenkamp (Dept. of Clinical Radiology, University of Münster). Gereon Heuft, Gudrun Schneider (Dept. of Psychosomatics and Psychotherapy, University of Münster). Thomas Reker (LWL-Hospital Münster). Gisela Bartling (IPP Münster). Ulrike Buhlmann (Dept. of Clinical Psychology, University of Münster).

Helene Dukal, Christine Hohmeyer, Lennard Stütz, Viola Schwerdt, Fabian Streit, Josef Frank, Lea Sirignano (Dept. of Genetic Epidemiology, Central Institute of Mental Health, Medical Faculty Mannheim, Heidelberg University). Stefanie Heilmann-Heimbach, Stefan Herms, Per Hoffmann (Institute of Human Genetics, University of Bonn, School of Medicine & University Hospital Bonn). Andreas J. Forstner (Institute of Human Genetics, University of Bonn, School of Medicine & University Hospital Bonn; Centre for Human Genetics, Marburg University).

Anastasia Benedyk, Miriam Bopp, Roman Keßler, Maximilian Lückel, Verena Schuster, Christoph Vogelbacher (Dept. of Psychiatry, Marburg University). Jens Sommer, (Core-Facility Brainimaging, Marburg University). Thomas W.D. Möbius (Institute of Medical Informatics and Statistics, Kiel University).

Julian Glandorf, Fabian Kormann, Arif Alkan, Fatana Wedi, Lea Henning, Alena Renker, Karina Schneider, Elisabeth Folwarczny, Dana Stenzel, Kai Wenk, Felix Picard, Alexandra Fischer, Sandra Blumenau, Beate Kleb, Doris Finholdt, Elisabeth Kinder, Tamara Wüst, Elvira Przypadlo, Corinna Brehm (Comprehensive Biomaterial Bank Marburg, Marburg University).

We are further deeply indebted to all participants of this study.

